# Choroid Plexus Cysts on 7T MRI Differentiate NMOSD from MS

**DOI:** 10.1101/2025.10.06.25337455

**Authors:** Zhiming Zhen, Siyao Xu, Can Xu, Yunyun Duan, Yicheng Hsu, Peiyu Huang, Yaou Liu, Li Gui, Chen Liu

## Abstract

**Background:** Multiple sclerosis (MS) and neuromyelitis optica spectrum disorder (NMOSD) share overlapping clinical and imaging features, complicating differential diagnosis. The choroid plexus is increasingly recognized as a regulator of neuroinflammation and may play a critical role in the pathogenesis and differentiation of these conditions. Choroid plexus cysts (CPCs) are difficult to detect with conventional MRI resolution and standard anatomical approaches. High-resolution 7T MRI enables their visualization as readily identifiable and quantifiable morphological changes, offering a new direction for choroid plexus research.

**Objective:** To characterize choroid plexus cysts (CPCs) in aquaporin-4 antibody–positive NMOSD and relapsing–remitting MS (RRMS) using ultra–high-field 7T MRI.

**Methods:** Fourteen patients aged 16–30 years were prospectively recruited, including seven NMOSD (mean age 22.9 years) and seven MS (mean age 22.1 years). CPCs were assessed on UHR-T2WI-TSE images for number, maximum/minimum diameter, and cross-sectional area. Conventional brain lesions were evaluated on T2-FLAIR. Independent readings by two radiologists were adjudicated by a senior neuroradiologist.

**Results:** CPCs were present in all NMOSD patients (7/7) but only 57% of MS patients (4/7). Compared with MS, NMOSD showed a higher CPC burden with greater counts (median 4 vs 1; *p*=0.037) and non-significant trends toward larger diameters and areas (maximum diameter 4.17 mm vs 2.27 mm; minimum diameter 3.58 mm vs 1.82 mm; cross-sectional area 4.41 mm^2^ vs 2.02 mm^2^; *p*=0.072–0.095). Both groups demonstrated right-sided predominance. In contrast, brain lesions were more prevalent in MS (7/7) than in NMOSD (3/7; *p*=0.015).

**Conclusions:** 7T MRI reveals distinct CPCs characteristics in NMOSD and MS. Despite fewer brain lesions in NMOSD compared with MS, CPCs burden remained consistently higher. These findings suggest that CPCs may represent a more sensitive and quantifiable structural change, serving as a potential imaging-biomarker for distinguishing NMOSD from MS. Validation in larger cohorts is warranted to advance understanding of neuroimmunological diseases and to clarify the role of CPCs in neuroinflammation.

**Key point:**

1. Identifying additional differences in brain injury between NMOSD and MS is crucial for diagnosis and therapy.
2. CPCs are more prevalent and distinct in NMOSD than in MS, unveiling an “iceberg” of structural pathology hidden at conventional imaging resolution.
3. CPCs are readily identifiable and quantifiable, indicating their potential as a novel imaging biomarker for differential diagnosis in clinical practice.

## Background

Neuromyelitis optica spectrum disorder (NMOSD) and multiple sclerosis (MS) are inflammatory demyelinating diseases that share clinical features but possess differing pathological mechanisms and require distinct optimal treatments.^1, 2^ Although oligoclonal bands and AQP4 antibodies provide supportive diagnostic value, they are not absolute criteria for differentiation. MRI is an essential tool for diagnosis and disease monitoring, with abnormalities such as hyperintense white matter lesions and lesion enhancement commonly seen in both disorders. Typical diagnostic MRI features include periventricular Dawson’s fingers in MS and involvement of AQP4-rich regions (e.g., the area postrema) in NMOSD. ^1, 2^ However, some patients may lack characteristic brain lesions or the requisite evidence of dissemination in time and space, and in certain cases, NMOSD patients may present with “MS-like” lesion patterns on brain MRI. Such imaging overlap complicates early and accurate diagnosis. ^1, 2^ It is critical to investigate new imaging biomarkers of these diseases for understanding potential pathogenesis, differential diagnosis, and patient management.

Glymphatic system dysfunction is closely associated with sustained central nervous system (CNS) inflammation and progressive tissue damage in neuroimmunological diseases. Within this system, the choroid plexus (CP) plays a pivotal role as a key structure, contributing to CSF production, metabolic regulation, immune cell trafficking, and the maintenance of CNS homeostasis. Substantial evidence indicates that CP is actively involved in the neuroinflammatory processes of MS.^3-5^ 3T MRI studies have demonstrated that enlarged CP volume (CPV) in MS correlates with white matter lesion burden, brain atrophy, higher relapse rate, and disability progression.^6-9^ In contrast, no significant CPV enlargement in NMOSD, suggesting that CPV may serve as a potential imaging-marker to differentiate MS from NMOSD.^6^ However, the accurate measurement of CPV still relies on complex segmentation methods, which remain challenging in clinical practice. In addition, morphological alterations of the CP have yet to be fully elucidated.

With advances in MRI technology, thin-walled CP cysts (CPCs), structures maintained by rapid blood flow and hydrostatic pressure, can now be visualized in vivo at 7T MRI. Beyond CPV, CPCs represent a novel and more accessible marker of CP pathology, referred to as the’grape sign’.^10^ In this article, we aim to characterize CPCs in NMOSD and MS, hypothesizing that CPCs occur in both disorders but are more frequent in NMOSD due to AQP4-antibody–mediated astrocytopathy.

## Methods

### Participants

We prospectively recruited AQP4+ NMOSD and relapsing–remitting MS (RRMS) patients. AQP4+ NMOSD were diagnosed according to the 2015 International Panel guidelines.^1^ RRMS were diagnosed according to the 2017 McDonald criteria.^2^ To minimize age effects, only participants aged 16–30 years were included^10^. Demographic data, disease characteristics, and selected clinical assessments were recorded (table 1). This study was approved by the Ethics Committee of the First Affiliated Hospital of Army Medical University [(A)KY2025059].

**Table 1.**
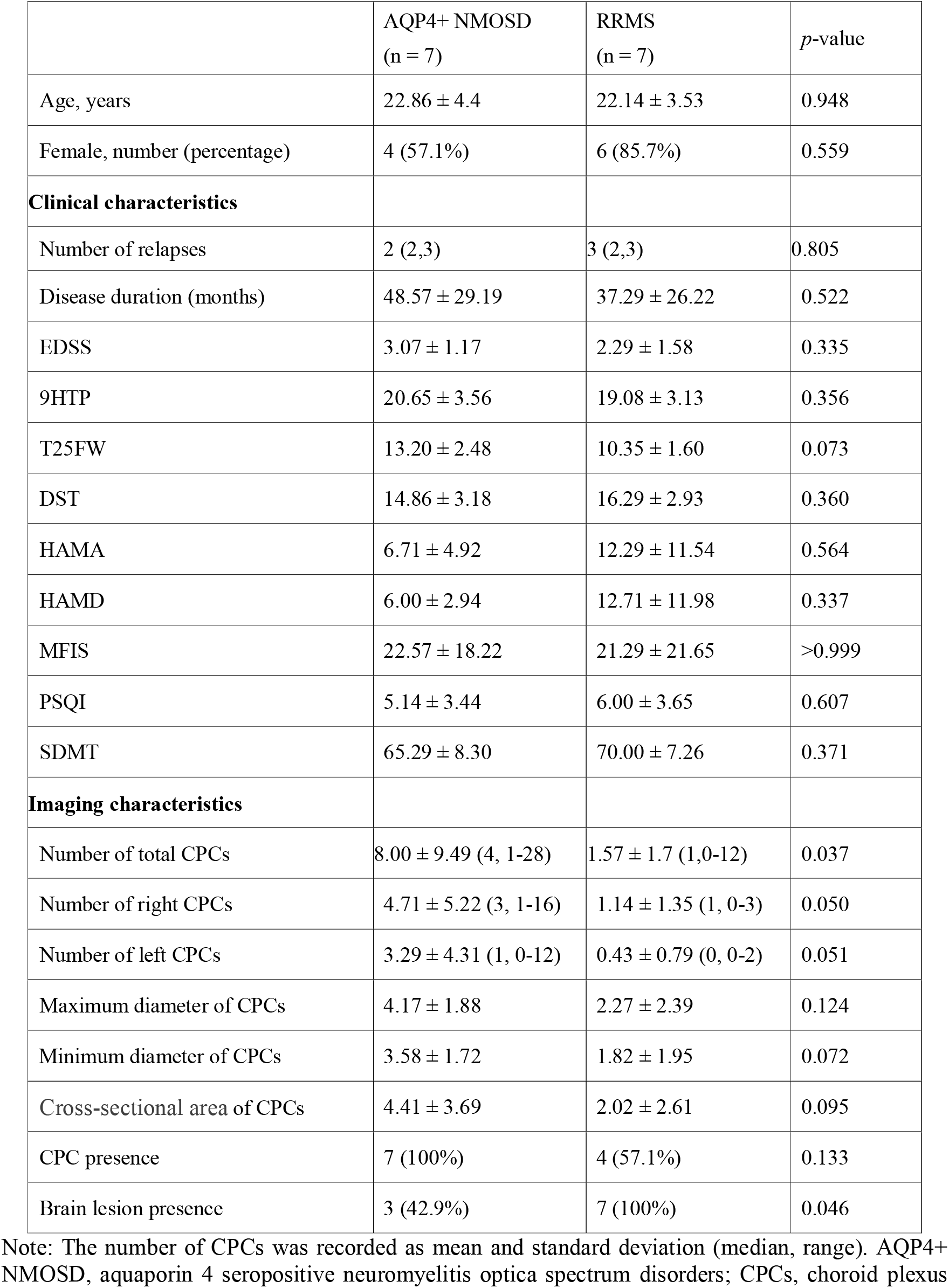
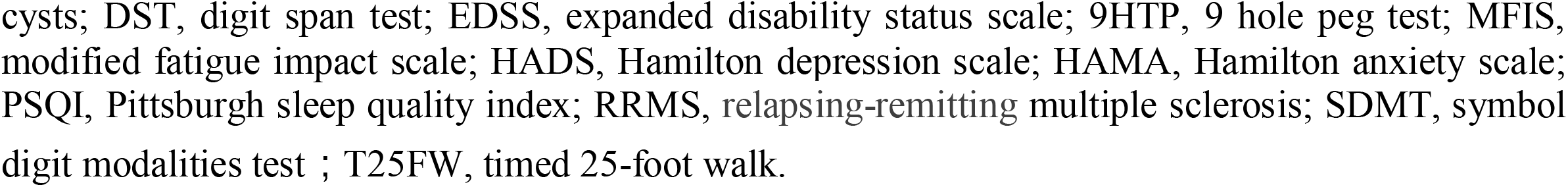
Demographic, clinical, and MRI characteristics.

|  | AQP4+ NMOSD<br>(n = 7) | RRMS<br>(n = 7) | *p*-value |
| --- | --- | --- | --- |
| Age, years | 22.86 ± 4.4 | 22.14 ± 3.53 | 0.948 |
| Female, number (percentage) | 4 (57.1%) | 6 (85.7%) | 0.559 |
| **Clinical characteristics** |  |  |  |
| Number of relapses | 2 (2,3) | 3 (2,3) | 0.805 |
| Disease duration (months) | 48.57 ± 29.19 | 37.29 ± 26.22 | 0.522 |
| EDSS | 3.07 ± 1.17 | 2.29 ± 1.58 | 0.335 |
| 9HTP | 20.65 ± 3.56 | 19.08 ± 3.13 | 0.356 |
| T25FW | 13.20 ± 2.48 | 10.35 ± 1.60 | 0.073 |
| DST | 14.86 ± 3.18 | 16.29 ± 2.93 | 0.360 |
| HAMA | 6.71 ± 4.92 | 12.29 ± 11.54 | 0.564 |
| HAMD | 6.00 ± 2.94 | 12.71 ± 11.98 | 0.337 |
| MFIS | 22.57 ± 18.22 | 21.29 ± 21.65 | >0.999 |
| PSQI | 5.14 ± 3.44 | 6.00 ± 3.65 | 0.607 |
| SDMT | 65.29 ± 8.30 | 70.00 ± 7.26 | 0.371 |
| **Imaging characteristics** |  |  |  |
| Number of total CPCs | 8.00 ± 9.49 (4, 1-28) | 1.57 ± 1.7 (1,0-12) | 0.037 |
| Number of right CPCs | 4.71 ± 5.22 (3, 1-16) | 1.14 ± 1.35 (1, 0-3) | 0.050 |
| Number of left CPCs | 3.29 ± 4.31 (1, 0-12) | 0.43 ± 0.79 (0, 0-2) | 0.051 |
| Maximum diameter of CPCs | 4.17 ± 1.88 | 2.27 ± 2.39 | 0.124 |
| Minimum diameter of CPCs | 3.58 ± 1.72 | 1.82 ± 1.95 | 0.072 |
| Cross-sectional area of CPCs | 4.41 ± 3.69 | 2.02 ± 2.61 | 0.095 |
| CPC presence | 7 (100%) | 4 (57.1%) | 0.133 |
| Brain lesion presence | 3 (42.9%) | 7 (100%) | 0.046 |

Note: The number of CPCs was recorded as mean and standard deviation (median, range). AQP4+ NMOSD, aquaporin 4 seropositive neuromyelitis optica spectrum disorders; CPCs, choroid plexus
cysts; DST, digit span test; EDSS, expanded disability status scale; 9HPT, 9 hole peg test; MFIS, modified fatigue impact scale; HADS, Hamilton depression scale; HAMA, Hamilton anxiety scale; PSQI, Pittsburgh sleep quality index; RRMS, relapsing-remitting multiple sclerosis; SDMT, symbol digit modalities test ; T25FW, timed 25-foot walk.

### MRI Acquisition and Analysis

All participants underwent 7T MRI (MAGNETOM Terra, Siemens) with a 32-channel head coil, including T2-weighted fluid-attenuated inversion recovery (T2-FLAIR) sequence (0.8 mm^3^-isotropy resolution), T1-weighted magnetization-prepared two rapid gradient-echo (T1-MP2RAGE) sequence (0.65mm^3^-isotropy resolution), ultra-high-resolution T2-weighted turbo spin-echo imaging (UHR-T2WI-TSE) sequence (0.2×0.2×1 mm^3^ resolution).

CPCs were defined as circular or oval cystic structures visible across two consecutive slices, with hypointense cyst walls and hyperintense internal content on T2WI (Figure 1) ^10^. Both T1-MP2RAGE and T2-FLAIR sequences showed low signal intensity within the cysts. Two radiologists (R.Z. and S.Y., each with 5 years of neuroimaging experience) independently assessed the number of bilateral CPCs on UHR-T2WI-TSE images and evaluated brain lesion locations on T2-FLAIR. The largest cyst was measured for its long- and short-axis diameters and cross-sectional area. Discrepancies were resolved by consensus with a senior neuroradiologist (C.L., 10 years of experience).

**Figure 1.**
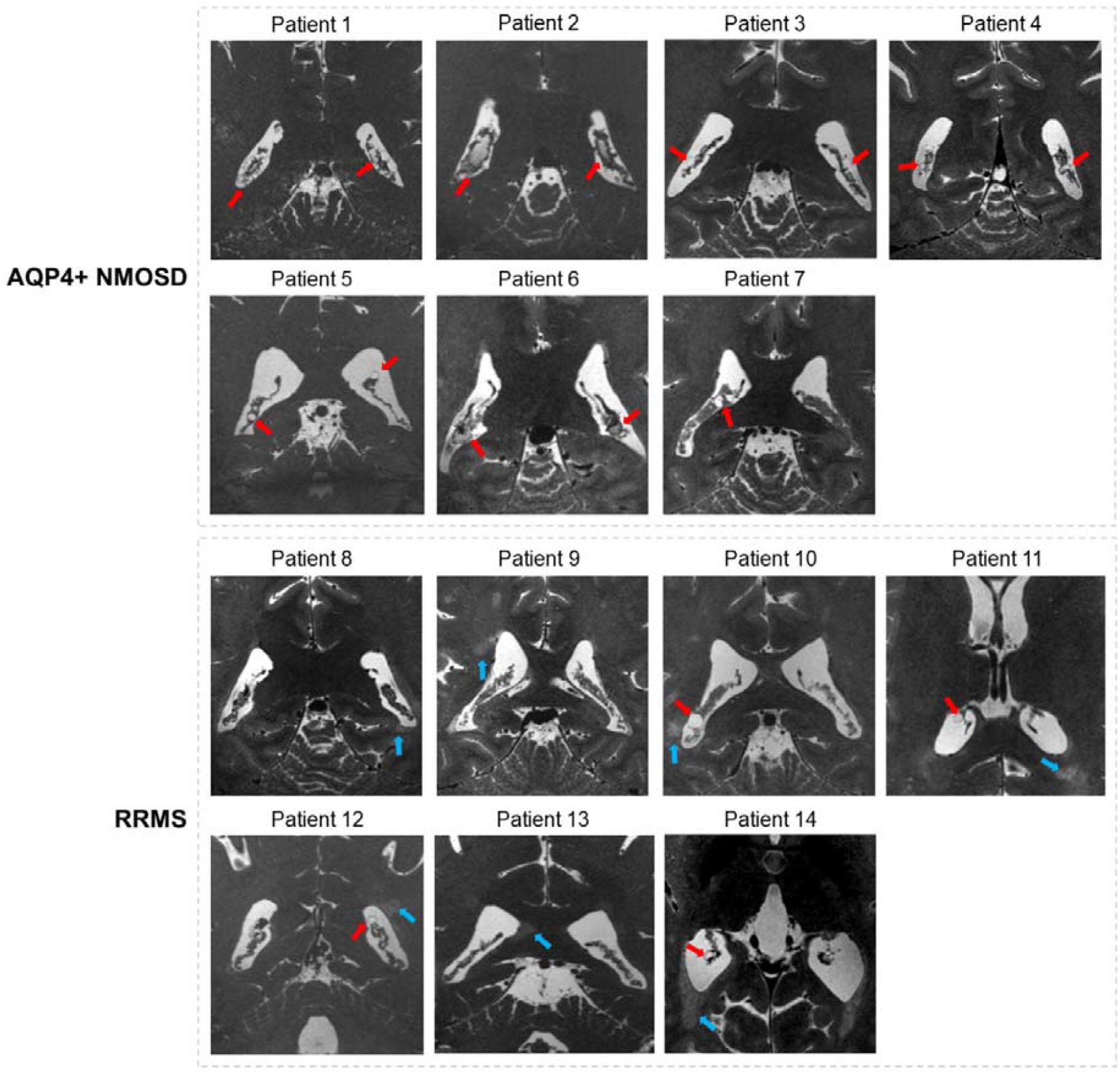
7T MRI UHR-T2WI-TSE images of choroid plexus cysts (CPCs) in AQP4+ NMOSD and RRMS patients. CPCs (red arrows) were observed in four RRMS patients and in all AQP4+ NMOSD patients. In NMOSD, CPCs showed more distinctive features, including greater number and larger size. All MS patients exhibited brain imaging abnormalities (blue arrows). AQP4+ NMOSD, aquaporin-4 seropositive neuromyelitis optica spectrum disorder; RRMS, relapsing-remitting multiple sclerosis.

### Statistical Analysis

Statistical analyses were performed using SPSS 22.0. Categorical variables were presented as numbers (percentages). Continuous variables were presented as mean ± standard deviation (SD). Ranked data were recorded as median and interquartile range. CPCs counts were shown as mean ± SD (median, range). Between-group analyses employed parametric t-tests, nonparametric Mann-Whitney U tests, or Chi-squared tests as appropriate, with statistical significance defined as *p*<0.05.

## Results

We initially recruited 9 AQP4+ NMOSD patients and 10 RRMS patients. However, 2 NMOSD cases and 3 MS cases were excluded due to motion artifacts and incomplete clinical scales. Ultimately, we enrolled 7 NMOSD patients (4 female, mean age 22.86 ± 4.4 years) and 7 RRMS patients (6 female, mean age 22.14 ± 3.53 years), with detailed clinical characteristics provided in Table 1.

As illustrated in Figure 1, NMOSD patients demonstrated higher CPC counts than MS (median 4 [1-28] vs 1 [0-4]; *p*=0.037), with right-sided predominance observed in both groups. CPC size parameters (diameters and cross-sectional area) tended to be larger in NMOSD but did not reach statistical significance (*p*=0.072–0.095).Notably, all NMOSD patients (7/7) exhibited CPCs compared to only 43% (3/7) showing brain lesions, while MS patients demonstrated the inverse pattern with 100% (7/7) brain lesion prevalence but only 57% (4/7) CPC occurrence (*p*=0.015). Table 2 presents case-specific CPC features with brain lesion location and clinical information.

**Table 2.** CPC imaging features, brain lesion location, and clinical characteristics by case.

|  | Number of total CPCs | Number of right CPCs | Number of left CPCs | Maximum diameter of total CPCss (mm) | Minimum diameter of total CPCs (mm) | Cross-sectional area of CPCs (mm) | Visible brain lesions on imaging | Number of relapses | Disease duration (months) | ED SS | SD MT |
| --- | --- | --- | --- | --- | --- | --- | --- | --- | --- | --- | --- |
| Patient 1 NMOSD | 28 | 16 | 12 | 5.48 | 4.97 | 6.8 | NA | 2 | 36 | 3 | 60 |
| Patient 2 NMOSD | 5 | 3 | 2 | 6.71 | 6.24 | 10 | Deep WM | 2 | 12 | 3.5 | 63 |
| Patient 3 NMOSD | 1 | 1 | 0 | 3.74 | 2.63 | 2.5 | Periependymal surfaces of fourth ventricle | 5 | 72 | 2.5 | 51 |
| Patient 4 NMOSD | 3 | 2 | 1 | 2.35 | 2.15 | 1.3 | DeepWM; Periventricular WM | 3 | 95 | 4.5 | 70 |
| Patient 5 NMOSD | 3 | 2 | 1 | 2 | 1.65 | 0.8 | NA | 3 | 44 | 1.5 | 69 |
| Patient 6 NMOSD | 4 | 3 | 1 | 5.98 | 4.7 | 7 | NA | 2 | 21 | 4.5 | 67 |
| Patient 7 NMOSD | 12 | 6 | 6 | 2.96 | 2.73 | 2 | NA | 2 | 60 | 2 | 77 |
| Patient 8 RRMS | 0 | 0 | 0 | 0 | 0 | 0 | Deep WM; Periventricular WM | 2 | 40 | 1 | 65 |
| Patient 9 RRMS | 0 | 0 | 0 | 0 | 0 | 0 | Deep WM; Periventricular WM; Cortical/juxtacortical | 3 | 60 | 0 | 72 |
| Patient 10 | 3 | 3 | 0 | 5.16 | 4.65 | 6 | Deep WM; | 3 | 18 | 3.5 | 59 |
| RRMS |  |  |  |  |  |  | Periventricular WM;<br>Cortical/juxtacortical |  |  |  |  |
| Patient 11<br>RRMS | 1 | 1 | 0 | 2.28 | 2.09 | 1.2 | Deep WM;<br>Periventricular WM;<br>Cortical/juxtacortical | 4 | 77 | 4 | 75 |
| Patient 12<br>RRMS | 3 | 1 | 2 | 5.44 | 4.02 | 5.5 | Cortical/juxtacortical | 2 | 7 | 4 | 80 |
| Patient 13<br>RRMS | 0 | 0 | 0 | 0 | 0 | 0 | Cortical/juxtacortical | 3 | 12 | 1.5 | 74 |
| Patient 14<br>RRMS | 4 | 3 | 1 | 3 | 2 | 1.5 | Deep WM;<br>Periventricular WM;<br>Cortical/juxtacortical | 2 | 47 | 2 | 65 |

Note: AQP4+NMOSD, aquaporin 4 seropositive neuromyelitis optica spectrum disorders; EDSS, expanded disability status scale; RRMS, relapsing-remitting multiple sclerosis; SDMT, symbol digit modalities test; WM, white matter.

## Discussion

To our knowledge, this is the first study to observe and characterize CPCs in patients with NMOSD and MS, offering a potential new perspective on CP involvement in these conditions. Our findings can be summarized in two major points: 1.CPCs were more prevalent in AQP4+ NMOSD than in RRMS on 7T MRI. Importantly, 7T imaging allows CPCs to be readily identified and quantified, revealing an ‘iceberg’ of structural pathology that remains concealed at conventional imaging resolution. 2. In AQP4+ NMOSD, CPCs were present even in patients without visible brain lesions on imaging, whereas in MS, patients with extensive parenchymal damage exhibited no CPCs. This highlights the potential of CPCs as a sensitive marker of CP involvement beyond conventional lesion assessment.

Previous studies focusing on CPV changes have demonstrated greater CP enlargement in MS compared to NMOSD.^6 11, 12^ However, CPCs remain invisible on 3T MRI CPV-imaging, as the limited contrast and resolution of structural imaging (1 mm^3^-ISO) and partial volume effects preclude reliable depiction of their thin-walled structure. In post-mortem anatomical and animal preparations, CPCs are prone to collapse, and reports of CPCs in neuroimmunological diseases have been scarce. 7T MRI overcomes these limitations, enabling clear visualization of CPCs beyond the conventional concept of CPV (Figure 2). Moreover, as a novel in vivo imaging finding, CPCs are easier for clinicians to identify in routine practice than CPV, which requires segmentation.

**Figure 2.**
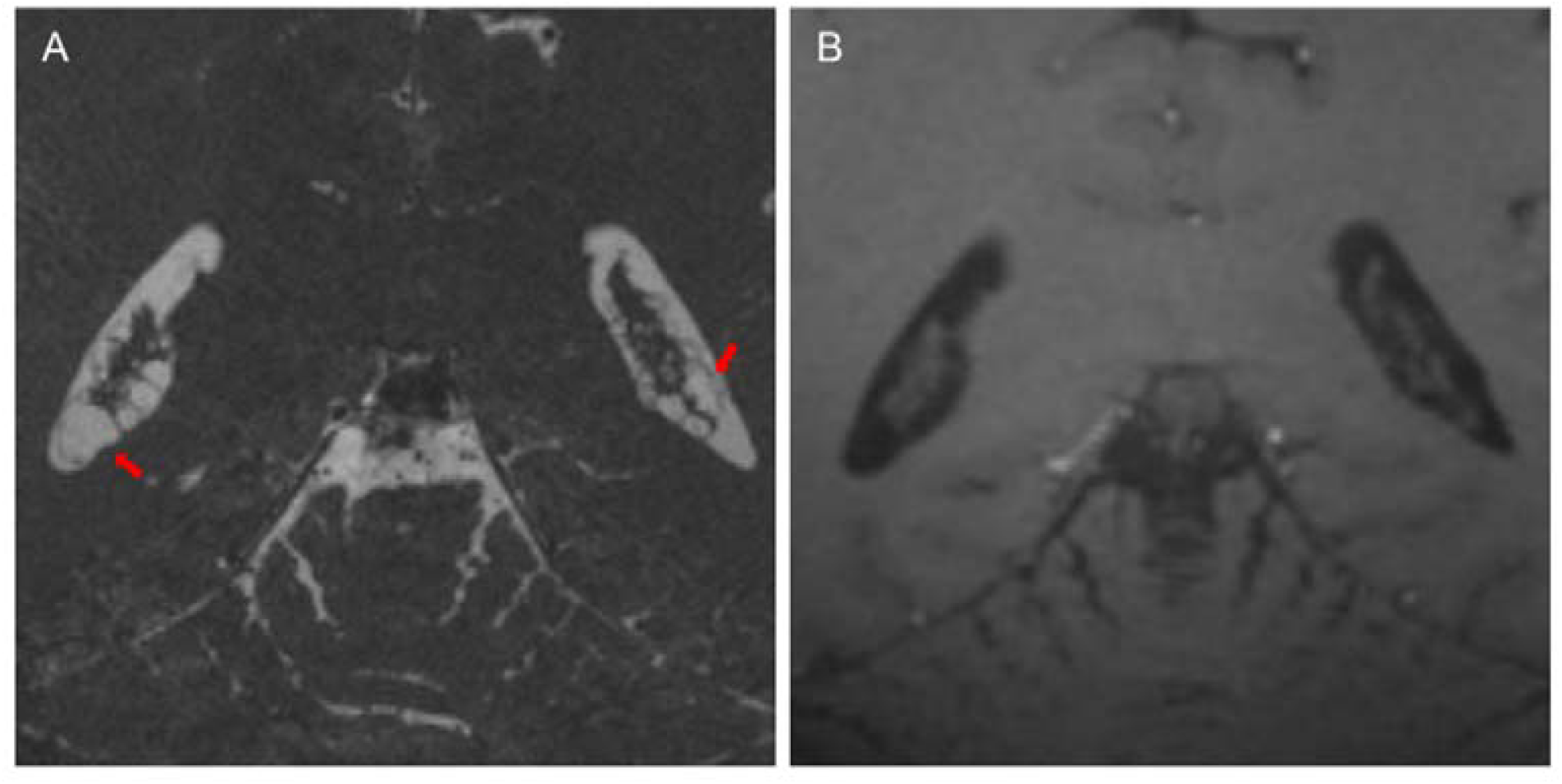
Comparison of 7T MR imaging of choroid plexus cysts (CPC). On the same imaging plane of the NMOSD patient, CPCs (red arrow) are visible with high-resolution UHR-T2WI-TSE (A) (0.2×0.2×1 mm^3^ resolution), whereas they are not visible on T1-MP2RAGE imaging (B) (0.65mm^3^-isotropy resolution). NMOSD, neuromyelitis optica spectrum disorders; T1-MP2RAGE, T1-weighted magnetization-prepared two rapid gradient-echo sequence; UHR-T2WI-TSE, ultra-high-resolution T2-weighted turbo spin-echo imaging sequence.

MS and NMOSD are neuroinflammatory disorders with distinct immunopathological targets. In MS, demyelination results primarily from immune-mediated injury to oligodendrocytes and myelin sheaths. Choroid plexus (CP) alterations in this context are considered secondary, with CPV enlargement reflecting immune cell trafficking, cytokine-driven edema, and stromal proliferation.^13,14^ In contrast, the involvement of the CP in NMOSD appears to be more direct. NMOSD is characterized by AQP4–IgG–mediated astrocytic injury, leading to complement activation and subsequent inflammatory demyelination. Recent studies have confirmed AQP4 expression in CP epithelium, providing a plausible anatomical substrate for more pronounced structural injury through direct antibody-mediated attac^3, 15^ Within this mechanistic framework, our findings support the hypothesis that antibody-mediated damage to AQP4-rich CP structures predisposes NMOSD patients to more frequent formation of CPCs compared with MS. These lesions may represent a more direct and therefore more sensitive imaging marker of CP involvement, detectable even in patients without overt parenchymal brain lesions.

In conclusion, our study demonstrates that CPCs are more prevalent in NMOSD than in MS on 7T MRI. This structural observation provides novel insights into CP involvement in these disorders and suggests that CPCs may represent a promising new imaging biomarker for differential diagnosis.

## Supporting information

STROBE

## Data Availability

The analyzed data in our study are subject to the following licenses/restrictions: The original anonymized imaging data analyzed in this manuscript will be used for research purposes. Requests for access to these datasets should be directly addressed to corresponding author.

## Acknowledgements

We extend our gratitude to the 7T Magnetic Resonance Imaging Translational Medical Center, Department of Radiology, Southwest Hospital, Third Military Medical University (Army Medical University) for their invaluable support in this study. A heartfelt thank you to the many screening participants and Chinese Neuroimmunology Patient Advocacy Organizations: Demyelinating Disease Patients’ Home, Multiple Sclerosis Home who provided significant insights from their experiences, benefitting others. We deeply cherish their generous contributions.

## Financial Disclosures of all Authors for the Past Year

None. We have not received any financial support, grants, or funding from commercial entities or organizations. We have not submitted this manuscript to multiple journals simultaneously.

## Ethical statement

The study was approved by the Ethics Committee of the First Affili-ated Hospital of the Army Medical University (KY2025059). Before the study, written informed consent was obtained from all participants.

## Author contributions

Guarantors of integrity of entire study, C.L., L.G.,Y.L; study concepts/study design or data acquisition or data analysis/interpretation, all authors; manuscript drafting or manuscript revision for important intellectual content,all authors; approval of final version of submitted manuscript, all authors; agrees to ensure any questions related to the work are appropriately resolved, all authors; literature research, Z.Z., S.X., D.D., P.H., Y.L., ; clinical studies, Z.Z., S.X., L.G., C.X.,; statistical analysis, Z.Z., S.X., C.X., Y.H.; and manuscript editing, Z.Z., S.X., C.L., L.G., Y.L;

